# Childhood trajectories of emotional and behavioral difficulties are related to polygenic liability for mood and anxiety disorders

**DOI:** 10.1101/2023.11.21.23298804

**Authors:** Nora Refsum Bakken, Nadine Parker, Laurie J. Hannigan, Espen Hagen, Pravesh Parekh, Alexey Shadrin, Piotr Jaholkowski, Evgeniia Frei, Viktoria Birkenæs, Guy Hindley, Laura Hegemann, Elizabeth C. Corfield, Martin Tesli, Alexandra Havdahl, Ole A. Andreassen

## Abstract

**Background:** Symptoms related to mood and anxiety disorders often present in childhood and adolescence. Some of the genetic liability for mental disorders, and emotional and behavioral difficulties seems to be shared. Yet, it is unclear how genetic liability for mood and anxiety disorders influence trajectories of childhood emotional and behavioral difficulties, and if specific developmental patterns associate with higher genetic liability for these disorders.

**Methods:** This study uses data from a genotyped sample of children (*n* = 54,839) from the Norwegian Mother, Father and Child Cohort Study (MoBa). We use latent growth models (1.5-5 years) and latent profile analyses (1.5-8 years) to quantify childhood trajectories and profiles of emotional and behavioral difficulties and diagnoses. We examine associations between these trajectories and profiles with polygenic scores for bipolar disorder (PGS_BD_), anxiety (PGS_ANX_), depression (PGS_DEP_), and neuroticism (PGS_NEUR_).

**Results:** Associations between PGS_DEP_, PGS_ANX_ and PGS_NEUR_, and emotional and behavioral difficulties in childhood were developmentally stable rather than age specific. Higher PGS_ANX_ and PGS_DEP_ were associated with steeper increases in behavioral difficulties across early childhood. Latent profile analyses identified five profiles. All PGS were associated with probability of classification into profiles characterized by some form of difficulties (vs. a normative reference profile), but only PGS_BD_ was uniquely associated with a single developmental profile.

**Conclusions:** Our findings indicate that genetic risk for mood disorders and related traits contribute to a more rapidly increasing and higher overall burden of emotional and behavioral difficulties across early and middle childhood, with some indications for disorder-specific profiles. These findings of associations between childhood trajectories and symptom profiles and genetic and clinical susceptibility for mental disorders, may form the basis for more targeted early interventions.

## INTRODUCTION

Mood and anxiety disorders are common and account for a large global burden of disease (Vos et al., 2020), but the disease mechanisms remain largely unknown (Bennett, 2023). The disorders are regarded as multifactorial, influenced by multiple genes in combination with lifestyle and environmental factors (Penninx, Pine, Holmes, & Reif, 2021; Malhi & Mann, 2018; Thapar, Eyre, Patel, & Brent, 2022). Twin and family studies have revealed significant heritability for mood and anxiety disorders, estimated at 60-90% for bipolar disorders (O’Connell & Coombes, 2021), 30-50% for major depressive disorder (Kendall et al., 2021) and 20-60% for anxiety disorders (Ask et al., 2021). Recent genome-wide association studies (GWAS) have shown that common genetic variants account for a substantial part of their heritability (Howard et al., 2019; Purves et al., 2020; Mullins et al., 2021), confirming a polygenic architecture (Andreassen, Hindley, Frei, & Smeland, 2023). The discoveries from GWAS have made it possible to derive polygenic scores (PGS) that capture an individual’s genetic liability to these disorders.

Childhood is a critical period in development of mood and anxiety disorders. Studies in high risk populations show that children of parents with an affective disorder are more likely to struggle with mental health issues such as emotional and behavioral problems than those without this family history (Lau et al., 2018; Rasic, Hajek, Alda, & Uher, 2013; Biederman et al., 2004; Weissman et al., 2005). Furthermore, population studies suggest that PGS for mood disorders and related traits are associated with childhood internalizing symptoms, ADHD symptoms and social problems (Akingbuwa et al., 2020; Akingbuwa, Hammerschlag, Bartels, & Middeldorp, 2022). However, how genetic liability is linked to developmental trajectories of emotional and behavioral difficulties is unclear (Moyakhe, Dalvie, Mufford, Stein, & Koen, 2023). Better characterization of this relationship is key to understand not only if, but how and when, genetic liability for mood and anxiety disorders manifest during childhood development.

Defining robust patterns of mental traits during childhood that may predict mood and anxiety disorders is needed to advance treatment options and prevention strategies (Hickie et al., 2013). Despite large advancements during the past decades, treatment options for mental disorders in general, and mood and anxiety disorders in spesific, are limited (Cuijpers, Stringaris, & Wolpert, 2020). Only 38% of young people receiving treatment for depression and/or anxiety in routine specialist mental health care report a reliable improvement (Bear, Edbrooke-Childs, Norton, Krause, & Wolpert, 2020), and earlier onset is associated with risk of a more severe course of disorder (Zisook et al., 2007; Joslyn, Hawes, Hunt, & Mitchell, 2016; Ramsawh, Weisberg, Dyck, Stout & Keller, 2011). This highlights the need for advanced tailored prevention and intervention initiatives targeting youth where current treatment effects are low and consequences severe. High genetic risk for mood and anxiety disorders have been assossiated with treatment resistance (Fanelli et al., 2022; Foo et al., 2019; Ward et al., 2018), earlier onset of disorder, and higher symptom-burden (Kwong et al., 2021; Wray et al., 2018). Thus, a better understanding of the genetic risk for mood and anxiety disorders in relation to trajectories of emotional and behavioral difficulties in childhood can inform development of more targeted preventive interventions.

In the current study we aim to identify how polygenic liability for depression (Howard et al., 2019), anxiety (Purves et al., 2020), bipolar disorder (Mullins et al., 2021), and neuroticism (Nagel et al., 2018) associate with developmental trajectories and profiles of childhood emotional and behavioral difficulties (measured at 1.5-8 years of age). We evaluate if these associations are developmentally stable or age specific, and if PGS mainly influence the overall level of emotional and behavioral difficulties or the rate of change in difficulties across childhood. Lastly, we investigate if higher genetic liabilities are associated with the likelihood of being assigned to specific developmental profiles and how this further is related to risk of emotional (mood or anxiety) disorders. We expect that these results will contribute to a deeper understanding of how polygenic liability is involved in the development of mood and anxiety disorders.

## METHODS

### Study population

The Norwegian Mother, Father and Child Cohort Study (MoBa) is a population-based pregnancy cohort study conducted by the Norwegian Institute of Public Health (NIPH). Participants were recruited from across Norway during the recruitment period of 1999 to 2008. The women consented to participation in 41% of the pregnancies (Magnus et al., 2016; Magnus et al., 2006). Blood samples obtained from the children’s umbilical cord at birth were used for genotyping (Paltiel et al., 2014). The current study is based on version 12 of the quality-assured maternally filled-out questionnaires released for research in January 2019. Out of the overall sample of 113,530 children, we selected a subset (*n* = 54,839) based on the following criteria: available quality controlled (Corfield et al., 2022) genotyping information, coupling to the Medical Birth Registry of Norway (MBRN) and pruning for genetic relatedness using KING version 2.2.5 (Manichaikul et al., 2010). The unrelated function (Manichaikul et al., 2012) was used to extract the maximal list of individuals that contains no first- or second-degree relatives (kinship coefficient threshold < 0.0885). For an overview of the complete sample selection process see Figure S1.

The establishment of MoBa and initial data collection was based on a license from the Norwegian Data Protection Agency and approval from The Regional Committees for Medical and Health Research Ethics (REC). MoBa follows regulations from the Norwegian Health Registry Act. The current study was approved by the administrative board of MoBa led by NIPH and REC (2016/1226/REK Sør-Øst C).

### Childhood emotional and behavioral difficulties

Measures of emotional and behavioral difficulties were based on questionnaires filled out by the mothers when their children were 1.5, 3, 5, and 8 years old. Emotional and behavioral difficulties in early childhood (1.5, 3, and 5 years) were measured using items from the corresponding subscales of The Child Behavior Checklist (CBCL) that were consistent across the three time points (repeated measures) (Achenbach & Ruffle, 2000). At the age of 8 years (middle childhood), more comprehensive measures of emotional and behavioral difficulties were used: a 13-item Short Mood and Feelings Questionnaire (SMFQ) measures depressive symptoms (Angold, Costello, Messer & Pickles, 1995), a 5-item short form of the Screen for Child Anxiety Related Disorders (SCARED) (Birmaher et al., 1999) measures symptoms of anxiety, and a 34-item Rating Scale for Disruptive Behavior Disorders (RS-DBD) (Silva et al., 2005) measures symptoms of conduct disorders (8 items), oppositional defiant disorders (8 items), hyperactivity (9 items), and inattention (9 items). The R-package “Phenotools v0.2.7” (Hannigan et al., 2023) was used to compute sum scores from the individual items. To be included in the computation of a sum score, at least half the subscales’ items were required to be non-missing. An overview of the individual items included in each measure is presented in Table S1.

### Emotional (mood or anxiety) disorder

Information on diagnoses were retrieved from the Norwegian Patient Registry (NPR), where International Classification of Diseases Tenth Revision (ICD-10) (World Health Organization, 2004) diagnoses registered in specialist health care services from 2008 through 2022 were available. As anxiety and mood disorders commonly co-occur and have overlapping symptomatology, we examined the disorders as a combined category of emotional disorders optimizing utility for the purpose of prevention and early detection (Bullis, Boettcher, Sauer-Zavala, Farchione & Barlow, 2019). The emotional disorders category comprised mood disorders (F30-F39), anxiety disorders (F40-F41), and emotional disorders with onset occurring in childhood and adolescence (F92-F93).

### Covariates

Given known sex differences in the prevalence of emotional disorders and related traits, and the wide range in birth years in MoBa, we included sex and birth year (categorized to three levels) as covariates in all analyses. To account for population stratification effects, the first ten genetic principal components (PCs) and genotyping batch were also included as covariates.

### GWAS summary statistics

This study relied on summary statistics for common emotional disorders and traits that were acquired from the largest GWAS on bipolar disorder (Mullins et al., 2020), anxiety (Purves et al., 2020), depression (Howard et al., 2019) and neuroticism (Nagel et al., 2018). As the bipolar disorder GWAS included MoBa study participants, summary statistics excluding these participants were used. The GWAS for neuroticism was computed on the UK Biobank genotypic samples.

### Statistical analyses

#### Polygenic scores (PGS)

We used LDpred2 (Privé, Arbel & Vilhjálmsson, 2021) to compute PGS_BD,_ PGS_ANX_, PGS_DEP,_ and PGS_NEUR,_ using their respective GWAS summary statistics and the correlation matrix between genetic variants (Privé, 2022). To account for population stratification and batch effects, we performed a linear regression with the standardized PGS as outcome variables (from LDpred2) and the first ten genetic principal components (PCs) and dummy-coded genotyping batches (26 batches). We used the standardized residuals from these models for further analyses (henceforth referred to as PGS). For detailed description of PGS-calculation see Supplementary text1. PGS relation to corresponding trait in our sample are provided in Table S2.

### Linear regression models (Cross-sectional associations)

We performed linear regression analyses to study the association between each measure of childhood emotional and behavioral difficulties and the polygenic scores. Separate linear regression models were fit for each of the measures of emotional and behavioral difficulties with the independent variables being PGS_ANX_, PGS_DEP,_ PGS_BD_ and PGS_NEUR_ (considered one at a time). To account for multiple testing, we performed a false discovery rate (Benjamini & Hochberg, 1995) correction for the 48 multiple comparisons (12 emotional and behavioral difficulties, four PGS). Prior to running these linear regression models, we standardized each of the 12 measures to have zero mean and unit standard deviations.

### Latent Growth Models (Developmental trajectories in early childhood)

Linear latent growth models were used to assess the influence of polygenic scores on developmental trajectories of emotional and behavioral difficulties across early childhood. First, we defined the latent growth models to model the developmental trajectories of emotional and behavioral difficulties across early childhood. We looked at Comparative Fit Index (CFI), Tucker Lewis Index (TLI) and Root Mean Square Error of Approximation (RMSEA), to evaluate if a linear growth model was adequate. Next, to assess the association between PGS and these developmental trajectories, we specified five models having different effect of PGS on the developmental trajectories: PGS effect on age specific residuals (PGS effects are *age specific*), PGS effect on intercept, slope, or both growth factors (PGS effects are *developmentally stable*; three models), and PGS effect fixed to null (PGS effects are *not present*). These models were compared in a “stepwise” manner using change in Akaike Information Criterion (AIC) and *p*-values from Chi square test at α = 0.05, choosing the least complex model when we observed non-significant differences (see Supplementary text 2 for model specification and Supplementary text 3 for details on this stepwise procedure). At the end of this procedure, we selected eight models (four PGS for two trajectories of emotional and behavioral difficulties). To account for multiple comparisons, we applied a false discovery rate (Benjamini & Hochberg, 1995) correction for the number of PGS effects identified across these eight models. The growth models were constructed using scripts adapted from (Hannigan et al., 2021), and were ran with the R package *lavaan* version 0.6-7 (Rossel, 2012).

### Latent Profile Analyses (Developmental profiles across early and middle childhood)

To extract characteristic developmental patterns of emotional and behavioral difficulties across early and middle childhood (age 1.5, 3, 5, and 8 years), we used latent profile analyses. We adapted latent profile models from our previous study (Bakken et al., 2023) and additionally included PGS_ANX_, PGS_DEP_, PGS_BD_ and PGS_NEUR_ as covariates. Here a 3-step maximum likelihood approach (Nylund-Gibson, Grimm & Masyn, 2019; Vermunt, 2010) were used to generate and assign profile membership to each individual. Specifically, for each individual, we used their estimated scores on the continuous latent growth factors (from the growth models), scores on emotional and behavioral difficulties at 8 years of age, emotional disorder diagnosis (distal outcome to validate profiles against clinical disorders), and PGS (as covariates; each PGS was used separately) (see Figure S2). Missing data were handled by full information maximum likelihood (FIML). We specified models with 2 to 8 profiles and selected the model (i.e., the number of profiles) by loglikelihood value (LL), first-order AIC, second-order AIC, Vuong–Lo– Mendell–Rubin likelihood ratio test (VLMR), entropy, the substantive interpretability of each profile, and the proportion of individuals assigned to the smallest profile (we used a threshold of > 1%) (Bakken et al., 2023; Petersen, Qualter & Humphrey, 2019). All analyses were conducted in Mplus version 8.3 (Muthén & Muthén, 1998–2017) and R version 4.0.3 leveraging the package “MplusAutomation” version 1.0.0 (Hallquist & Wiley, 2018).

## RESULTS

Demographic and descriptive information for key study variables is presented in Table 1. A flow diagram illustrating sample population at each stage of analysis is presented in Figure S1.

**Table 1.** Demographic characteristics and descriptive information on key study variables.

|  | **Emotional**<br>**disorder**<br>(*n* = 4, 968) | **No emotional**<br>**disorder**<br>(*n* = 49, 871) | **Total**<br>(*n* = 54, 839) |
| --- | --- | --- | --- |
| Sex, male, *n* (%) | 1827 (36.78%) | 26281 (52.70%) | 28108 (51.26%) |
| Age end of follow-up, years, mean (SD) | 17.60 (2.14) | 16.90 (2.15) | 16.97 (2.16) |
| Age first emotional disorder, years, mean, (SD) | 14.20 (3.54) | - | 14.20 (3.54) |
| Highly educated mother or father, *n* (%) | 2749 (63.55%) | 32636 (73.21%) | 35385 (72.35%) |
| Maternal age, mean, (SD)<sup>a</sup> | 29.76 (4.81) | 30.25 (4.59) | 30.21 (4.61) |
| Parity, *n* (%) primiparous | 22652 (45.42%) | 2227 (44.83%) | 24879 (45.37%) |
| Mother's country of birth |  |  |  |
| Norway, *n* (%) | 4611 (94.70%) | 46318 (94.48%) | 50929 (94.50%) |
| Other high-income-country, *n* (%) | 205 (4.21%) | 2132 (4.35%) | 2337 (4.34%) |
| Other GDB 7 super region country, *n* (%) | 53 (1.09%) | 573 (1.17%) | 626 (1.16%) |
| Depressive disorder, *n* (%) | 1954 (39.33%) | 0 (0.00%) | 1954 (3.56%) |
| Anxiety disorder, *n* (%) | 2853 (57.43%) | 0 (0.00%) | 2853 (5.20%) |
| Bipolar disorder, *n* (%) | 118 (2.38%) | 0 (0.00%) | 118 (0.22%) |
| ADHD, *n* (%) | 1005 (20.23%) | 2676 (5.37%) | 3681 (6.71%) |
| Obsessive compulsive disorder, *n* (%) | 243 (4.89%) | 323 (0.65%) | 566 (1.03%) |
| Conduct disorder, *n* (%) | 102 (2.05%) | 251 (0.50%) | 353 (0.64%) |
| **Key study variables, mean, (SD)** |  |  |  |
| Emotional difficulties: |  |  |  |
| 1.5 years | 1.45 (1.29) | 1.30 (1.21) | 1.31 (1.21) |
| 3 years | 1.61 (1.52) | 1.39 (1.38) | 1.41 (1.40) |
| 5 years | 1.48 (1.59) | 1.05 (1.27) | 1.08 (1.30) |
| Behavioral difficulties: |  |  |  |
| 1.5 years | 4.10 (2.32) | 3.92 (2.25) | 3.94 (2.25) |
| 3 years | 4.29 (2.60) | 3.83 (2.40) | 3.86 (2.42) |
| 5 years | 2.97 (2.54) | 2.45 (2.23) | 2.48 (2.26) |
| Emotional and behavioral difficulties<br>at age 8 years: |  |  |  |
| Depressive symptoms | 2.77 (3.14) | 1.78 (2.36) | 1.86 (2.44) |
| Anxiety symptoms | 1.44 (1.57) | 0.98 (1.14) | 1.02 (1.19) |
| Inattention | 5.80 (4.66) | 4.89 (4.06) | 4.96 (4.11) |
| Hyperactivity | 4.20 (4.54) | 3.52 (3.86) | 3.57 (3.92) |
| Conduct disorder symptoms | 0.98 (1.78) | 0.73 (1.46) | 0.75 (1.49) |
| Oppositional defiant disorder<br>symptoms | 4.33 (3.76) | 3.31 (3.06) | 3.39 (3.14) |

**Note:** ADHD, attention-deficit/hyperactivity disorder. a Mean maternal age at pregnancy is calculated for mothers between age 17 and 45 due to lack of information on age for mothers above or below this age-range.

### Cross-sectional associations between polygenic scores and emotional and behavioral difficulties in early and middle childhood

PGS for depression (PGS_DEP_), neuroticism (PGS_NEUR_) and anxiety (PGS_ANX_) were positively associated with all measures of emotional and behavioral difficulties across early and middle childhood (ages 1.5-8 years) (Figure 1 and Table S3-S5). PGS for bipolar disorder (PGS_BD_) was only significantly associated with depressive symptoms, and symptoms of oppositional defiant disorder, hyperactivity, and conduct disorder at 8 years (Figure 1 and Table S6).

**Figure 1.**
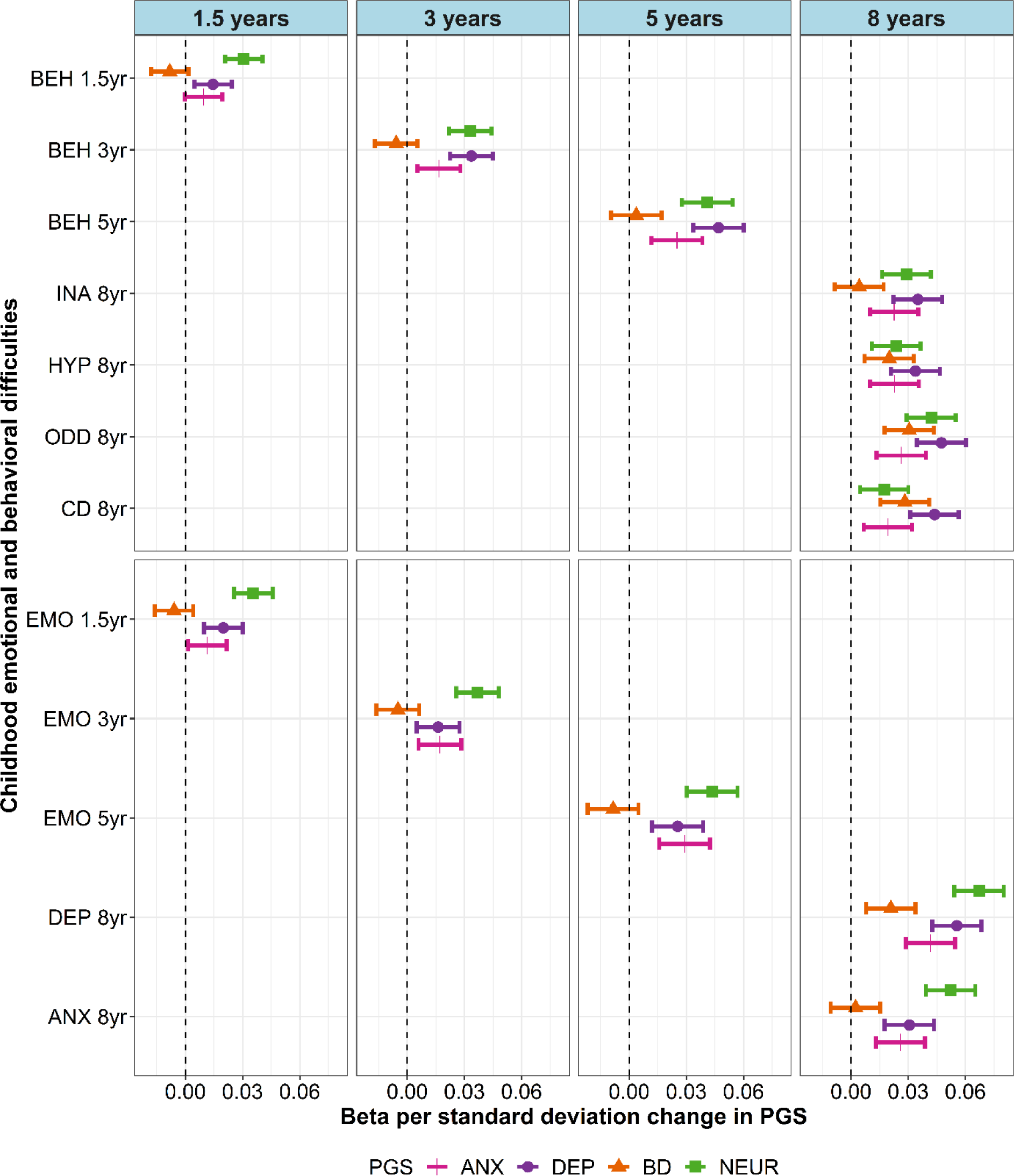
Cross-sectional associations between polygenic scores (PGS) and emotional and behavioral difficulties across early and middle childhood.

**Note**: *emo1.5yrs* = emotional difficulties at 1.5 years, *emo3yr* = emotional difficulties at 3 years, *emo5yr* = emotional difficulties at 5 years, *beh1.5yrs* = behavioral difficulties at 1.5 years, *beh3yr* = behavioral difficulties at 3 years, *beh5yr* = behavioral difficulties at 5 years, *dep8yr* = depressive symptoms at 8 years, *anx8yr* = anxiety symptoms at 8 years, *ina8yr* = inattention symptoms at 8 years, *odd8yr* = oppositional defiant disorder symptoms at 8 years, *cd8yr* = conduct disorder symptoms at 8 years. ANX = anxiety, DEP = depression, BD = bipolar disorder, NEUR = neuroticism.

### Polygenic scores association to developmental trajectories of emotional and behavioral difficulties in early childhood

The results from the latent growth models are presented in table S7-S11 and Table 2. Linear latent growth models provided a good fit for measures of emotional and behavioral difficulties across early childhood (CFI > 0.9, TIL>0.9, RMSEA<0.05) (Marsh et al., 2009) (Table S7). We found PGS_DEP_, PGS_ANX_ and PGS_NEUR_ to have overall stable associations, rather than age-specific effects, with emotional and behavioral difficulties across early childhood (1.5-5 years). We further found that for PGS_DEP_ the association with emotional difficulties were primarily related to overall levels of difficulties (βL_intercept_ = 0.029, 95% CI 0.018-0.041), while for behavioral difficulties PGS_DEP_ were related to both overall level and rate of change in difficulties (β_Dintercept_ = 0.021, 95% CI 0.009-0.034, βL_slope_ = 0.041, 95% CI 0.024-0.058). Higher PGS_ANX_ were related to a more rapid increase in emotional and behavioral difficulties (β_Dslope_ = 0.018, 95% CI 0.001-0.036 and β_Dslope_ = 0.018, 95% CI 0.001-0.035), and higher overall levels of emotional and behavioral difficulties (β_Dintercept_ = 0.015, 95% CI 0.002-0.029, βL_intercept_ = 0.013, 95% CI 0.001-0.029). PGS_NEUR_ were associated with higher overall level of emotional (β_Dintercept_ = 0.055, 95% CI 0.043-0.066) and behavioral difficulties (βL_intercept_ = 0.045, 95% CI 0.035-0.056). In accordance with the cross-sectional regression analyses, we did not find an effect of PGS_BD_ on the trajectories of emotional or behavioral difficulties in our latent growth models (the null effect model was no worse fitting than the simplest model including PGS_BD_ effects).

**Table 2.** Standardized beta for PGS on best performing latent growth model for emotional and behavioral difficulties across early childhood.

| Model | $\beta$ | 95% CI | *p*-value | *p*-FDR |
| --- | --- | --- | --- | --- |
| PGS<sub>DEP</sub> effect on intercept emotional difficulties | 0.029 | (0.018, 0.041) | <0.001 | <0.001 |
| PGS<sub>DEP</sub> effect on intercept behavioral difficulties | 0.021 | (0.009, 0.034) | 0.001 | 0.001 |
| PGS<sub>DEP</sub> effect on slope behavioral difficulties | 0.041 | (0.024, 0.058) | <0.001 | <0.001 |
| PGS<sub>NEUROT</sub> effect on intercept emotional difficulties | 0.055 | (0.043, 0.066) | <0.001 | <0.001 |
| PGS<sub>NEUROT</sub> effect on intercept behavioral difficulties | 0.045 | (0.035, 0.056) | <0.001 | <0.001 |
| No PGS<sub>BD</sub> effect on emotional difficulties | - | - | - |  |
| No PGS<sub>BD</sub> effect on behavioral difficulties | - | - | - |  |
| PGS<sub>ANX</sub> effect on intercept emotional difficulties | 0.015 | (0.002, 0.029) | 0.026 | 0.039 |
| PGS<sub>ANX</sub> effect on slope emotional difficulties | 0.018 | (0.001, 0.035) | 0.043 | 0.043 |
| PGS<sub>ANX</sub> effect on intercept behavioral difficulties | 0.013 | (0.001, 0.026) | 0.034 | 0.042 |
| PGS<sub>ANX</sub> effect on slope behavioral difficulties | 0.018 | (0.001, 0.035) | 0.037 | 0.042 |

**Note:** ANX = anxiety, DEP = depression, BD = bipolar disorder, NEUR = neuroticism. $\beta$ = Standardized Beta. 95% CI = 95% Confidence Interval. *p*-value = unadjusted *p*-value. $p\text{-FDR}$ = False discovery rate correction for multiple testing using Benjamini-Hochberg false discovery rate procedure.

### Developmental profiles of emotional and behavioral difficulties across early and middle childhood and the profiles relation to emotional disorders and polygenic scores

Based on LL, AIC, AICc, VLMR, entropy, distribution and the substantive interpretability of each profile we identified 5 developmental profiles of emotional and behavioral difficulties across early and middle childhood (Figure 2, Table S12-Table S13). The identified profiles were consistent with the profiles derived from the complete MoBa sample (Bakken et al., 2023).

**Figure 2.**
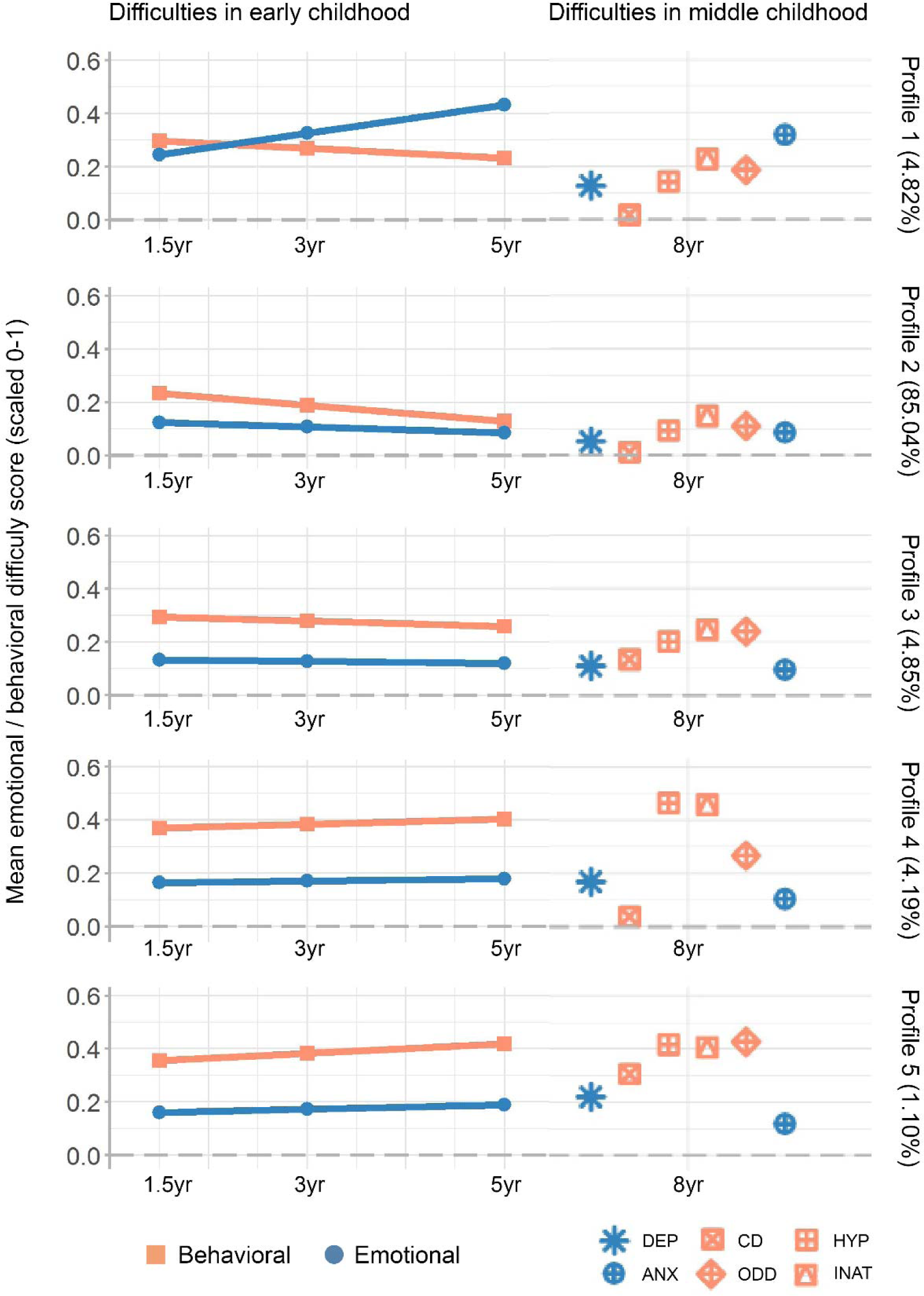
Developmental profiles of emotional and behavioral difficulties across early and middle childhood (1.5-8 years of age)

**Note**: Developmental profiles of childhood emotional and behavioral difficulties. Left panel shows development of early childhood emotional (blue, circle) and behavioral (red, square) difficulties measured by CBCL at 1.5 years, 3Lyears, and 5Lyears of age. Right panel shows corresponding middle childhood mental health trait with specific difficulty illustrated by shape at 8 years of age. DEP, depressive symptoms; CD, conduct disorder symptoms; HYP, hyperactivity symptoms; ANX, anxiety symptoms; ODD, oppositional defiant disorder symptoms; INAT, inattention symptoms.

**Profile 1 (4.82% of the sample)**: characterized by elevated and increasing emotional difficulties and slightly elevated behavioral difficulties across early childhood, and elevated levels of anxiety at the age of 8 years (“Anxiety profile”). **Profile 2 (85,04%):** characterized by low and decreasing levels of emotional and behavioral difficulties in early childhood (1.5-5 years), and low levels of emotional and behavioral difficulties at the age of 8 years (“Reference profile”).

**Profile 3 (4.85%):** characterized by slower decreasing behavioral symptoms across early childhood, and slightly elevated behavioral difficulties at the age of 8 years (“Delayed decreasing profile”). **Profile 4 (4.19%):** characterized by elevated levels of emotional and behavioral difficulties across early childhood, and elevated levels of hyperactivity, inattention and oppositional defiant disorder symptoms at 8 years (“ADHD profile”). **Profile 5 (1,10%)**: characterized by increasing and elevated behavioral and emotional difficulties across early childhood and elevated levels of depression, hyperactivity, inattention, oppositional defiant and conduct disorder symptoms at 8 years (“Behavioral dysregulation profile”).

Profiles 5 and 1, dominated by increasing and/or high levels of behavioral and emotional difficulties in childhood, had the strongest relationship with later diagnosis of emotional disorders (Table S14).

All PGS were associated with the likelihood of being assigned to a symptomatic profile rather than the reference profile (profile 2) in which symptoms were low and decreasing throughout childhood (Figure 3, Table S15-S18). Individuals with higher PGS_DEP_ were more likely to be assigned to profile 1 (“Anxiety”), 3 (“Delayed decreasing”), 4 (“ADHD”) or 5 (“Behavioral dysregulation”), compared to the reference profile (Figure 3, Table S15). Individuals with higher PGS_ANX_ were more likely to be assigned to profile 1 (“Anxiety”) or 3 (“Delayed decreasing”) compared to the reference profile (Figure 3, Table S16). Individuals with higher PGS_BD_ were most likely to be assigned to profile 5 (“Behavioral dysregulation”) (Figure 3, Table S17).

**Figure 3.**
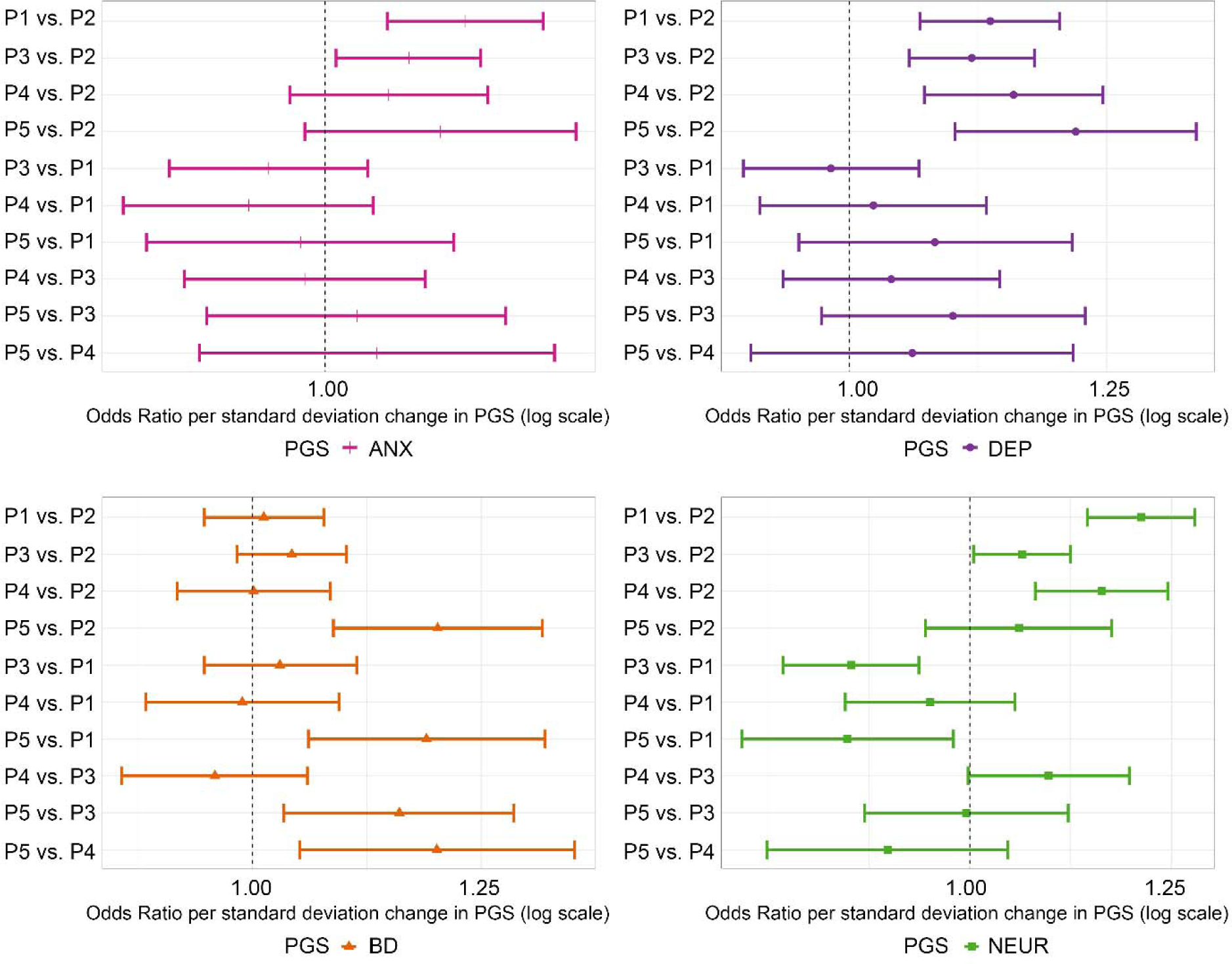
Relationship between polygenic scores and developmental profiles of emotional and behavioral difficulties across early and middle childhood (1.5-8 years of age)

**Note:** Odds Ratio comparing odds of being assigned to individual profiles per standard deviation increase in polygenic score (PGS) showed with 95% confidence intervals. ANX = anxiety, DEP = depression, BD = bipolar disorder, NEUR = neuroticism. P1 = Profile 1, P2 = Profile2, P3 = Profile 3, P4 = Profile 4, P5 = Profile 5.

Individuals with higher PGS_NEUR_ were more likely to be assigned to profile 1 (“Anxiety”), 3 (“Delayed decreasing”) or 4 (“ADHD”) compared to the reference profile. PGS_NEUR_ were more strongly associated with profile 1 (“Anxiety”), than profile 3 (“Delayed decreasing”) and profile 5 (“Behavioral dysregulation”), but no differences were found when compared to profile 4 (“ADHD”) (Figure 3, Table S18). No differences between odds of assignment to the different high-symptom-profiles (Profile 1, 3, 4 and 5) were found for PGS_DEP_ and PGS_ANX_ (Figure 3, Table S15-S16).

## DISCUSSION

The main findings of the current study were that genetic liability for mood and anxiety disorders is associated with overall higher and more rapidly increasing levels of emotional and behavioral difficulties across early and middle childhood, and that the genetic effect on developmental trajectories was mainly developmentally stable and less age specific. Furthermore, we found that PGS_DEP_ and PGS_ANX_ showed a general association with developmental profiles with higher levels of difficulties, while PGS_BD_ was specifically linked to a developmental profile characterized by elevated and increasing emotional and behavioral difficulties across early childhood, followed by elevated levels of depression and behavioral difficulties at 8 years of age (“Behavioral dysregulation”, profile 5). Overall, we find that children with higher genetic liability for mood and anxiety disorders are more likely to present with higher and/or more rapidly increasing levels of difficulties across childhood, difficulties that here and previously (Bakken et al., 2023) have been linked to likelihood of clinical disorder.

Our findings indicate that PGS_DEP_, PGS_ANX_ and PGS_NEUR_ show a developmentally stable, rather than age specific, association with emotional and behavioral difficulties. This is consistent with a cross-cohort meta-analysis showing no evidence of age as a moderator for association between PGS_DEP_ or PGS_NEUR_ in relation to childhood ADHD problems or internalizing problems (Akingbuwa et al., 2020), and association between PGS_SCZ_ and similar childhood measures as in the present study (Hannigan et al., 2021). The identified stability across age can be interpreted in several ways. First, stability across age could mean that genetic variants of importance to emotional and behavioral difficulties are expressed and involved in pathophysiological processes prior to onset of the difficulties (e.g in utero and infancy). It could also mean that the genetic variants collectively are involved with an approximately equal effect across the investigated developmental period (1.5-5 years). The presented explanations are neither fully encompassed, nor mutually exclusive, as both explanations can be true at the same time. Furthermore, as indicated from the cross-sectional analyses, the results from the latent growth models only show that the genetic effects are generally developmentally stable, not that there are no age specific effects at all. Such possibilities are coherent with findings of SNP heritability estimates varying with age (Akingbuwa et al., 2022), thus illustrating the importance of methodological consistency and the need for further studies replicating our findings in other samples and addressing the question of developmental stability with same and different methodology, and across a wider age span.

We find that polygenic liability for anxiety and depression are associated with a steeper increase and higher overall level of behavioral difficulties across early childhood. A genetically mediated acceleration in difficulties can be explained in several ways. First, these findings are coherent with theories of gene-environment correlations. Gene-environment correlations is the process in which an individual’s genotype influence, or is linked to, their exposure to environmental factors (Knafo & Jaffee, 2013). In this study we show that PGS_ANX_ and PGS_DEP_ are linked to higher levels of emotional and behavioral difficulties across childhood. The genetically linked elevated levels of difficulties could contribute to increased vulnerability for, or exposure to, environmental stressors such as: more negative attention from parents and/or other adult figures (Mcclowry et al., 2013), sensitivity to bullying, and problems with peer relationships (Perren, von Wyl, Stadelmann, Burgin & Von Klitzing, 2006; Klima & Repetti, 2008). Thus, further contributing to accelerated experience of emotional and behavioral difficulties, as identified in this study. Furthermore, neurobiological differences have been found to be associated with genetic risk for mood disorders and related traits (Alex et al., 2023; Shen et al., 2020; Fernandez-Cabello et al., 2022; Alnæs et al., 2019; Cheng et al., 2022; Patel et al., 2021), indicating potential for a neurodevelopmental component directly or indirectly contributing to higher and increasing emotional and behavioral difficulties, as well as risk of disorder.

From a clinical and public health perspective, our findings indicate that genetic information may help identify the children that are more likely to follow a less favorable trajectory of higher symptom burden across childhood. PGS_DEP,_ PGS_ANX_ and PGS_NEUR_ were all related to developmental profiles with higher levels of emotional and/or behavioral difficulties.

Furthermore, PGS_BD_ was specifically associated to profile 5 characterized by elevated and increasing behavioral symptoms, and high levels of depression, conduct disorder symptoms, oppositional defiant disorder symptoms, and hyperactivity and inattention at 8 years of age. The link between this developmental pattern and genetic risk for bipolar disorder, is consistent with studies finding CBCL-pediatric bipolar disorder phenotype (CBCL-PBD) and measures of emotional dysregulation in childhood to be related to later development of BD in adolescence and adulthood (Disalvo et al., 2023; Meyer et al., 2009; Biederman et al., 2009). Furthermore, in our analyses these results seem to be driven by a pattern of difficulties at 8 years of age. The lack of association between emotional and behavioral difficulties, and PGS_BD_ in early childhood is consistent with findings from smaller and/or cross-sectional studies (Biederman, Green, DiSalvo & Faraone, 2021; Duffy & Carlson, 2013; Akingbuwa et al., 2020). Our findings of a specific and general link between genetic liability and unfavorable developmental profiles of emotional and behavioral difficulties indicate that information on genetic risk may be an informative additional objective marker in risk evaluation of children presenting with mood-related symptoms. However, it is important to emphasize that currently, genetic risk as captured by PGS only explains a small amount of variation in symptom-burden. Therefore, further studies are needed to evaluate the utility of PGS in clinical samples and for individual-level predictions.

Strengths of the current study include a large number of individuals from a prospective population-based cohort, providing the opportunity to detect small effects, typical in complex phenotypes with polygenic architectures. Access to repeated measures of symptoms provide the opportunity to investigate both cross-sectional measures and trajectories. The longitudinal data enabled us to draw conclusions about relationships between genetic risk and symptoms across time. Nevertheless, there are also several limitations to consider. First, genetic liability as measured by PGS only captures relative risk based on additive effects of a subset of identified SNPs. Thus, non-additive effects and rare variants are not investigated, which may also influence the tendencies identified in this study (Kendall et al., 2019; Rees & Kirov, 2021). Second, PGSs are only available for individuals of European ancestry, thus reducing the generalizability to other ancestries. Further, the present measures of emotional and behavioral difficulties are based on questionnaires completed by mothers, which could bias the assessments. Last, the summary statistics from the GWAS are based on genetic risk for adult diagnoses. Some studies indicate that other variants may contribute to earlier onset of disorder (Kang et al., 2021; Power et al., 2017; Power et al., 2012; Harder et al., 2022), but this is still not well established. In the future, as GWAS on child and adolescents will become available, our findings should be replicated with PGS derived from child and adolescent diagnosis alone and combined with diagnosis at an older age.

## CONCLUSION

In conclusion, we find that polygenic liability for anxiety, depression, and neuroticism manifest in emotional and behavioral difficulties in early childhood with a developmentally stable rather than age specific effect. The genetic liability for anxiety and depression showed a general association with developmental profiles with higher levels of difficulties, while genetic risk for bipolar disorder were specifically related to symptoms consistent with behavioral dysregulation. Our results show that genetic risk for emotional disorders and related traits contribute to a higher overall burden and a faster increase in emotional and behavioral difficulties across early and middle childhood. These findings indicate presence of behavioral and emotional premorbid characteristics associated with genetic and clinical risk of disorder, thus suggesting a potential role of PGS in guiding targeted early interventions.

## ACKNOWLEDGEMENTS

We are grateful to all the participating families in Norway who take part in this on-going cohort study. We thank NIPH for generating high-quality genomic data. This research is part of the HARVEST collaboration, supported by the Research Council of Norway (RCN) (#229624). We also thank the NORMENT Centre for providing genotype data, funded by RCN (#223273), South East Norway Health Authorities and Stiftelsen Kristian Gerhard Jebsen. We further thank the Center for Diabetes Research, the University of Bergen (UiB) for providing genotype data and performing quality control and imputation of the data funded by the ERC AdG project SELECTionPREDISPOSED, Stiftelsen Kristian Gerhard Jebsen, Trond Mohn Foundation, RCN, the Novo Nordisk Foundation, UiB, and the Western Norway Health Authorities. We also thank deCODE genetics for providing the majority of the genotyping of the MoBa sample.

## DATA AVAILABILITY

Data from MoBa and MBRN used in this study are managed by the national health register holders in Norway (NIPH) and can be made available to researchers, provided approval from REC, compliance with the EU General Data Protection Regulation (GDPR) and approval from the data owners. The consent given by the participants does not open for storage of data on an individual level in repositories or journals. Researchers who want access to data sets for replication should apply through helsedata.no. Access to data sets requires approval from REC in Norway and an agreement with MoBa.

## KEY POINTS AND RELEVANCE

- There is a relationship between emotional and behavioral difficulties in childhood and genetic risk for mood and anxiety disorders.
- The association between genetic liability for anxiety, depression and neuroticism, and emotional and behavioral difficulties in childhood is developmentally stable. Genetic risk for depression and anxiety is associated with higher overall levels, and a more rapid increase, in emotional and behavioral difficulties during childhood.
- The trajectories of emotional and behavioral difficulties associated with genetic susceptibility for mental disorders, indicate potential opportunities for development of targeted early interventions.

