## Supporting information for "Childhood trajectories of emotional and behavioral difficulties are related to polygenic liability for mood and anxiety disorders"

### Figure S1 Flow diagram for study sample

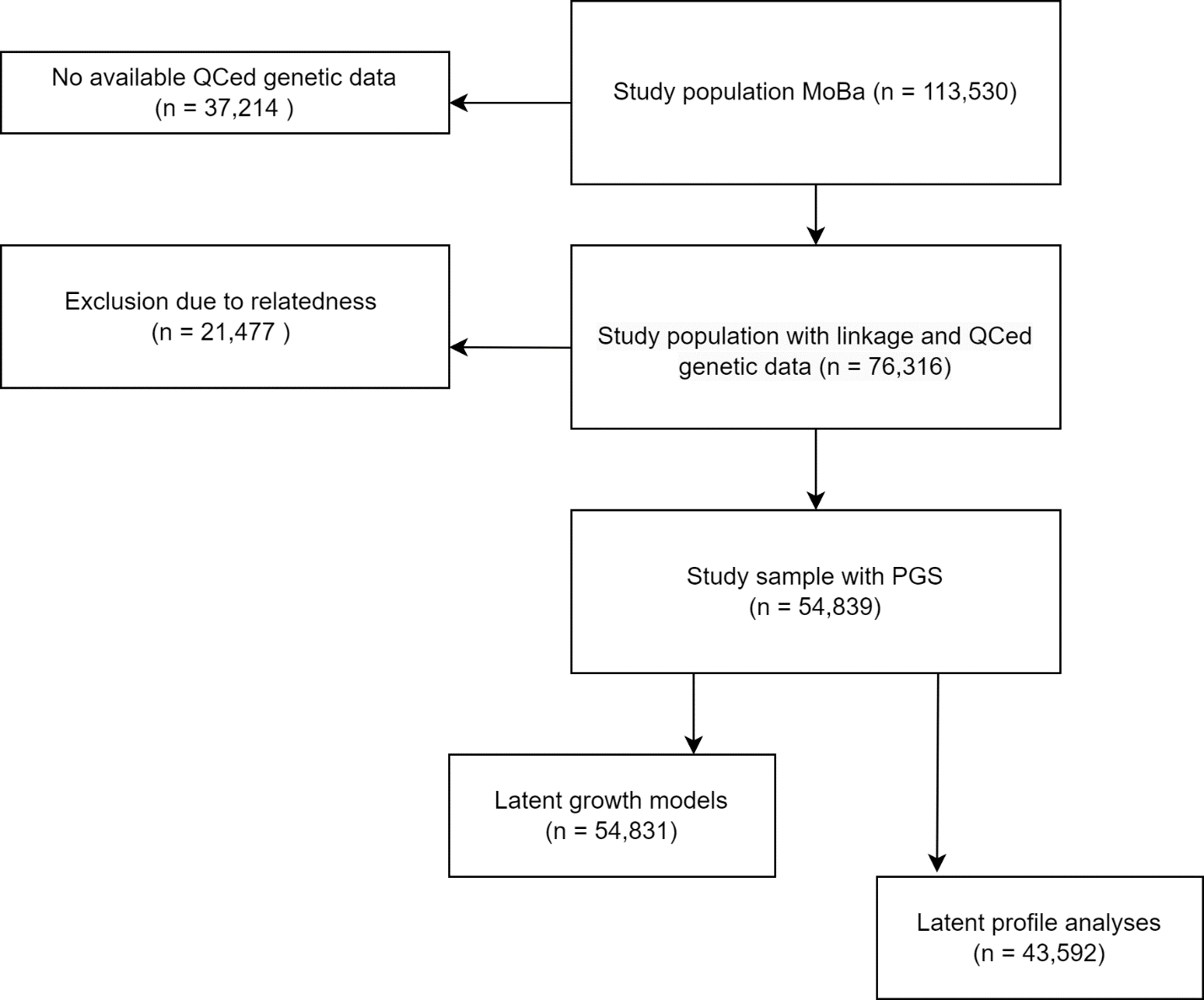

### Table S1 Questions included in each childhood emotional and behavioral difficulty measure.

| **Instrument** | **Mental health difficulty measures** | **Item level data for each measure** |
| --- | --- | --- |
| **CBCL** | **To what extent are the following statements true of your child’s behavior during the last two months?** | |
|  | Emotional difficulties | - Disturbed by any change in routine - Clings to adults or too dependent - Gets too upset when separated from parents - Too fearful or anxious - Doesn’t eat well |
|  | Behavioral difficulties | - Can’t concentrate, can’t pay attention for long - Cant sit still, restless or overactive - Hits others - Doesn’t seem to feel guilty after misbehaving - Gets in many fights - Quickly shifts from one activity to another - Defiant - Punishment doesn’t change his/her behaviour |
| **SMFQ** | **Mark how true each item has been for your child during the two last weeks.** | |
|  | Depressive symptoms 8yrs | - Felt miserable or unhappy - Felt so tired that s/he just sat around and did nothing - Was very restless - Didn’t enjoy anything at all - Felt s/he was no good anymore - Cried a lot - Hated him/herself - Thought s/he could never be as good as other kids - Felt lonely - Thought nobody really loved him/her - Felt s/he was a bad person - Felt s/he did everything wrong - Found it hard to think/concentrate |
| **SCARED** | **The questions below are about how your child have felt or behaved recently** | |
|  | Anxiety symptoms 8yrs | - My child gets really frightened for no reason at all - My child is afraid to be alone in the house - People tell my child that he/she worries too much - My child is scared to go to school - My child is shy |
| **RS-DBD** | **Mark the box that best describes your child’s behavior during the last 12 months/last year** | |
|  | Hyperactivity 8yrs | - Fidgets with hands or feet or squirms in seat (sits uneasily) - Leaves seat in classroom or in other situations in which remaining seated is expected (e.g. at the table or in group gathering) - Runs about or climbs excessively in situations in which it is inappropriate - Has difficulty playing or engaging in leisure activities quietly - Is “on the go” or acts as if “driven by a motor” - Talks excessively - Blurts out answers before questions have been completed - Has difficulty awaiting turn - Interrupts or intrudes on others, such as in conversation or play |
|  | Inattention 8yrs | - Fails to give close attention to details or makes careless mistakes in schoolwork - Has difficulty sustaining attention in tasks or play activities - Does not seem to listen when spoken to directly - Does not follow through on instructions and fails to finish school work, chores or duties (not due to oppositional behaviour or failure to understand instructions) - Has difficulty organizing tasks and activities - Avoids, dislikes or is reluctant to engage in tasks that require sustained mental effort (such as schoolwork or homework) - Loses things necessary for tasks or activities (pencils, books, toys) - Is easily distracted - Is forgetful in daily activities |
|  | Oppositional defiant disorder symptoms 8yrs | - Loses temper (tantrums) - Argues with adults - Actively defies or refuses to comply with adults’ requests or rules - Deliberately annoys people - Blames others for his/her mistakes or misbehaviour - Is touchy or easily annoyed by others - Is angry and resentful - Is spiteful or vindictive |
|  | Conduct disorder symptoms 8yrs | - Bullies, threatens or intimidates others - Initiates physical fights - Has been physically cruel to others - Has harassed or injured animals physically - Has stolen items of nontrivial value without confronting a victim (e.g. shoplifting) - Has deliberately destroyed other’s property - Has been truant from school - Has used an object that can cause serious physical harm to others (e.g. a bat, stone, knife, heavy toy) |

***Note:* CBCL=** The Child Behavior Checklist (Achenbach & Ruffle, 2000). **SMFQ=** Short Mood and Feelings Questionnaire (Angold, Costello, Messer, & Pickles, 1995). **SCARED=** Screen for Child Anxiety Related Disorders (Birmaher et al., 1999). **RS-DBD=** Rating Scale for Disruptive Behaviour Disorders (RS-DBD) (Silva et al., 2005).

### Supplementary text1

*Detailed description of PGS calculation using LDPred2*

SNP data from the unrelated children (*n* = 54,839) in MoBa were extracted and converted from PLINK .bed files to the bigsnpr R package “bigSNP” (Privé, Aschard, Ziyatdinov & Blum, 2018) format (using the bigsnpr::snp_readBed function), and imputed in-place (using bigsnpr::snp_fastImputeSimple function with mode “mean0”). Duplicate SNPs were subsequently removed. Polygenic scores were estimated using the LDPred2 “auto” model (Privé, Arbel & Vilhjálmsson, 2020), as implemented using the R package bigsnpr via a set of custom R scripts and software containers (Akdeniz, Frei, Hagen, Filiz, Karthikeyan, 2022; Frei, Jangmo, Hagen, Akdeniz, Zetterberg, Filiz & Shorter, 2023). The Ldpred2 “auto” model is free of hyperparameters. We used the LD reference for HapMAP3+ (Privé, 2022).

*PGS evaluation*

In order to evaluate the PGS for each trait (PGS_trait_), we fit generalized linear models (GLM) on the form

$$y_{\mathrm{trait}}\sim1+\mathrm{PGS}_{\mathrm{trait}}+SEX+\mathrm{BATCH}_{\mathrm{geno}}+\mathrm{Year}_{\mathrm{birth}}+\mathrm{PC}_{1}+\mathrm{PC}_{2}+\ldots+\mathrm{PC}_{10}.$$

Here, $y_{\mathrm{trait}}$ is the binary trait in terms of bipolar disorder (BD), anxiety (ANX), and depression (DEP), or neuroticism score (NEUR) which were treated as a continuous trait. The variable $\mathrm{SEX}$ denotes biological sex, $\mathrm{BATCH}_{\mathrm{geno}}$ the genotypic batch number, Year_birth_ the year of birth, and $\mathrm{PC}_{n}$ the $n$-th PC. SEX, BATCH_geno_ and Year_birth_ were treated as categorical variables (one-hot-encoded). The corresponding null model excluding effects of PGS_trait_ were fitted using GLMs on the form

$$y_{\mathrm{trait}}\sim1+SEX+\mathrm{BATCH}_{\mathrm{geno}}+\mathrm{Year}_{\mathrm{birth}}+\mathrm{PC}_{1}+\mathrm{PC}_{2}+\ldots+\mathrm{PC}_{10}.$$

The models were fitted using the “glm” function in R using the binomial family with logit links for the binary traits. The continuous trait models used the Gaussian family. Each logistic regression model’s goodness of fit was evaluated in terms of the Nagelkerke pseudo coefficient of determination (R^2^) (Nagelkerke, 1991), while the standard coefficient of determination (R^2^) was used for the continuous trait. The results are summarized in Table S2.

*Standardized residuals for PGS*

We calculated the residual PGS (for each trait) $\mathrm{PG}S_{\mathrm{trait}}^{\mathrm{res}}$ as:

$$\mathrm{PG}S_{\mathrm{trait}}^{\mathrm{res}}=PGS_{\mathrm{trait}}-X\hat{\beta},$$

where $\mathrm{PG}S_{\mathrm{trait}}$ is the standardized polygenic score from LDpred2, and $\hat{\beta}$ are the estimated linear regression coefficients obtained from regressing the effect of covariates $X$ (genotyping batch and the first ten genetic principal components) on $\mathrm{PG}S_{\mathrm{trait}}$:

$$\mathrm{PG}S_{\mathrm{trait}}\sim1+\mathrm{BATC}H_{\mathrm{geno}}+PC_{1}+\ldots+PC_{10}$$

### Table S2 PGS relation to each trait in our sample corresponding to trait from original GWAS

| **PGS** | **OR/**$\beta$ **(95% CI)** | ***p*-value** | **R^2^ PGS-model** | **R^2^ null model** |
| --- | --- | --- | --- | --- |
| **DEP** | 1.43 (1.36, 1.49) | <0.001 | 0.088 | 0.072 |
| **ANX** | 1.24 (1.19, 1.29) | <0.001 | 0.055 | 0.048 |
| **BD** | 1.58 (1.32, 1.90) | <0.001 | 0.078 | 0.063 |
| **NEUR** | 0.26 (0.21, 0.31) | <0.001 | 0.015 | 0.011 |

**Note:** Odds Ratio (OR) reported for diagnostic outcome. The standardized $\beta$ coefficient is reported for the continuous neuroticism trait. P-value corresponds to OR/$\beta$ for diagnostic outcome. PGS_DEP_ is tested against depressive disorder diagnosis (*n* = 1,954), PGS_ANX_ is tested against anxiety disorder diagnosis (*n* = 2,853), PGS_BD_ is tested against bipolar disorder diagnosis (*n* = 118) and PGS_NEUR_ is tested against maternally reported level of neuroticism at 8 years (continuous measure).

### Supplementary text 2

*Lavaan script for growth models*

#adapted from: <https://github.com/psychgen/scz-prs-psychopathol-dev/blob/master/scripts/01.1_specify_growth_models.R>

by_cat=birth year, pgs= polygenic score adjusted for 10 first principal components and batch effects.

**#Latent growth model without PGS to assess adequacy of linear model in sample**

basic <-

'

#Growth parameters (latent variables)

i1 =~ 1*ytime1 + 1*ytime2 + 1*ytime3

s1 =~ 0*ytime1 + 1.5*ytime2 + 3.5*ytime3

#Obs variable variances

ytime1 ~~ ytime1

ytime2 ~~ ytime2

ytime3 ~~ ytime3

#Growth parameter (co)variances

i1 ~~ i1

s1 ~~ s1

i1 ~~ s1

#Obs variable intercepts (fixed to 0)

ytime1 ~ 0*1 + sex + by_cat

ytime2 ~ 0*1 + sex + by_cat

ytime3 ~ 0*1 + sex + by_cat

#Growth parameter intercepts (freely estimated)

i1 ~ 1

s1 ~ 1

'

**# PGS effect on age specific residuals**

pgs_residuals <-

'

#Growth parameters (latent variables)

i1 =~ 1*ytime1 + 1*ytime2 + 1*ytime3

s1 =~ 0*ytime1 + 1.5*ytime2 + 3.5*ytime3

#Obs variable variances

ytime1 ~~ ytime1

ytime2 ~~ ytime2

ytime3 ~~ ytime3

#Growth parameter (co)variances

i1 ~~ i1

s1 ~~ s1

i1 ~~ s1

#Obs variable intercepts (fixed to 0)

ytime1 ~ 0*1 + pgs + sex + by_cat

ytime2 ~ 0*1 + pgs + sex + by_cat

ytime3 ~ 0*1 + pgs + sex + by_cat

#Growth parameter intercepts (freely estimated)

i1 ~ 1

s1 ~ 1

'

**# PGS on growth factors (intercept and slope)**

pgs_gf <-

'

#Growth parameters (latent variables)

i1 =~ 1*ytime1 + 1*ytime2 + 1*ytime3

s1 =~ 0*ytime1 + 1.5*ytime2 + 3.5*ytime3

#Obs variable variances

ytime1 ~~ ytime1

ytime2 ~~ ytime2

ytime3 ~~ ytime3

#Growth parameter (co)variances

i1 ~~ i1

s1 ~~ s1

i1 ~~ s1

#Obs variable intercepts (fixed to 0)

ytime1 ~ 0*1 + sex + by_cat

ytime2 ~ 0*1 + sex + by_cat

ytime3 ~ 0*1 + sex + by_cat

#Growth parameter intercepts (freely estimated)

i1 ~ 1 + pgs

s1 ~ 1 + pgs

'

**# PGS on intercept only**

pgs_intercept <-

'

#Growth parameters (latent variables)

i1 =~ 1*ytime1 + 1*ytime2 + 1*ytime3

s1 =~ 0*ytime1 + 1.5*ytime2 + 3.5*ytime3

#Obs variable variances

ytime1 ~~ ytime1

ytime2 ~~ ytime2

ytime3 ~~ ytime3

#Growth parameter (co)variances

i1 ~~ i1

s1 ~~ s1

i1 ~~ s1

#Obs variable intercepts (fixed to 0)

ytime1 ~ 0*1 + sex + by_cat

ytime2 ~ 0*1 + sex + by_cat

ytime3 ~ 0*1 + sex + by_cat

#Growth parameter intercepts (freely estimated)

i1 ~ 1 + pgs

s1 ~ 1 + 0*pgs

'

**# PGS effect on slope only**

pgs_slope <-

'

#Growth parameters (latent variables)

i1 =~ 1*ytime1 + 1*ytime2 + 1*ytime3

s1 =~ 0*ytime1 + 1.5*ytime2 + 3.5*ytime3

#Obs variable variances

ytime1 ~~ ytime1

ytime2 ~~ ytime2

ytime3 ~~ ytime3

#Growth parameter (co)variances

i1 ~~ i1

s1 ~~ s1

i1 ~~ s1

#Obs variable intercepts (fixed to 0)

ytime1 ~ 0*1 + sex + by_cat

ytime2 ~ 0*1 + sex + by_cat

ytime3 ~ 0*1 + sex + by_cat

#Growth parameter intercepts (freely estimated)

i1 ~ 1 + 0*pgs

s1 ~ 1 + pgs

'

**# no PGS effects (PGS fixed to null)**

lgm_nopgs <-

'

#Growth parameters (latent variables)

i1 =~ 1*ytime1 + 1*ytime2 + 1*ytime3

s1 =~ 0*ytime1 + 1.5*ytime2 + 3.5*ytime3

#Obs variable variances

ytime1 ~~ ytime1

ytime2 ~~ ytime2

ytime3 ~~ ytime3

#Growth parameter (co)variances

i1 ~~ i1

s1 ~~ s1

i1 ~~ s1

#Obs variable intercepts (fixed to 0)

ytime1 ~ 0*1 + sex + by_cat

ytime2 ~ 0*1 + sex + by_cat

ytime3 ~ 0*1 + sex + by_cat

#Growth parameter intercepts (freely estimated)

i1 ~ 1 + 0*pgs

s1 ~ 1 + 0*pgs

'

**Supplementary text 3**

*Detailed description of stepwise model selection procedure*

1. Define the five models: PGS effect on age specific residuals (“pgs_residuals”), PGS effect on both intercept and slope growth factors (“pgs_gf”), PGS effect on intercept growth factor (“pgs_intercept”), PGS effect on slope growth factor (“pgs_slope”), and PGS effect fixed to null (“lgm_nopgs).
2. Compare pgs_gf with pgs_residuals; if pgs_gf is not worse performing than pgs_residuals, select pgs_gf and move to step 3; otherwise select pgs_residuals as the final model.
3. If pgs_gf is selected at step 2, formally compare to pgs_intercept and pgs_slope respectively; if both are significantly worse performing, select pgs_gf as the final model; otherwise select the model with the lower AIC from pgs_intercept and pgs_slope and move to step 4.
4. If pgs_intercept or pgs_slope is selected at step 3, compare the selected model with lgm_nopgs; if lgm_nopgs is not worse performing, select it as the final model; otherwise, select the comparator model (either pgs_intercept or pgs_slope) as the final model.

We used the change in AIC and *p* value from Chi-square test to assess if the models were not worse performing.

### Figure S2: Latent profile analysis incorporating latent growth models for developmental profiles of emotional and behavioral difficulties and polygenic scores.

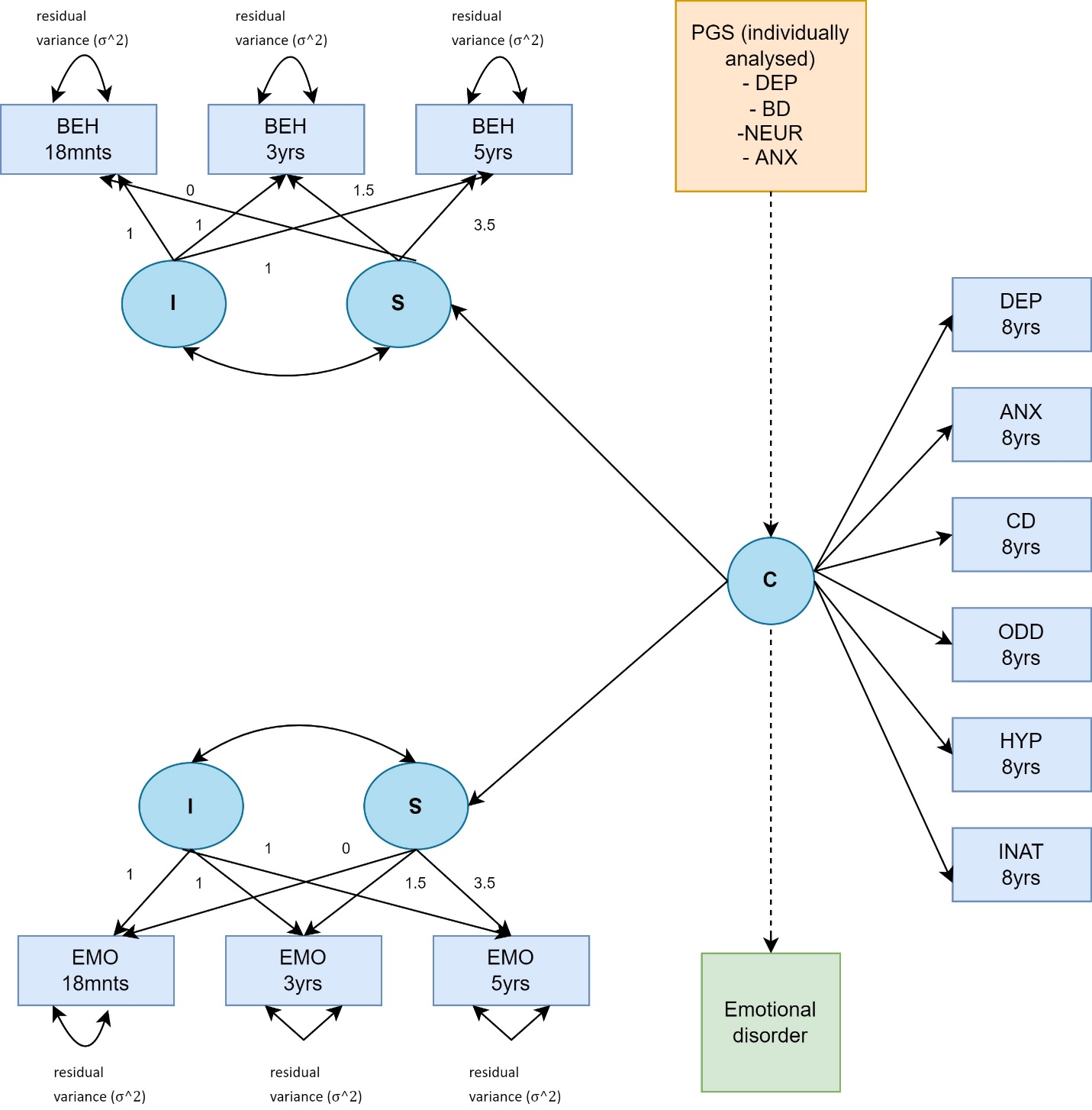

**Note**: Illustration of methods for developmental profiles incorporating latent growth models in 3-step maximum likelihood latent profile analysis. Boxes represent observed variables and circles model-estimated latent variables; I = intercept factor, loads equally on observed variables at all waves; C = categorical latent variable, subdividing the sample into a specified number of classes according to values: S = slope factor with loadings 0(18mnts), 1.5(3yrs), and 3.5(5yrs), corresponding to temporal distance between the measurements; BEH = Behavioral difficulties; EMO = Emotional difficulties; DEP= Depressive symptoms; ANX = Anxiety symptoms; CD= Conduct disorder symptoms; ODD = Oppositional defiant disorder symptoms; HYP = Hyperactivity; INAT= Inattention. Emotional/ behavioral intercept/slope variables and 8-year observed variables are intercorrelated within class (paths omitted from diagram for clarity) in step 1 of the latent profile analysis. Polygenic score for depression, anxiety, bipolar disorder and neuroticism is included as covariates (separately) and emotional disorder is included as distal outcome in step 3 in model-specification (indicated by dashed line).

For further methodological details on latent growth models see “Genetic liability for schizophrenia and childhood psychopathology in the general population” by L.J. Hannigan in Schizophrenia Bulletin Volume 47, Issue 4, July 2021, Pages 1179–1189 (Hannigan, et al., 2021) and “Childhood temperamental, emotional, and behavioral characteristics associated with mood and anxiety disorders in adolescence: A prospective study” by N.R. Bakken in Acta Psychiatrica Scandinavica Volume 147, Issue 2, February 2023. Pages 217-228 (Bakken, et.al, 2023).

Table S3: Depression PGS. Linear association between standardized PGS score and standardized score of emotional and behavioral difficulties.

| Characteristic | *n* | $\beta$ | 95% CI^1^ | *p*-value | *p-FDR*^2^ |
| --- | --- | --- | --- | --- | --- |
| Emotional difficulties 1.5 yrs | 36,984 | 0.02 | 0.01, 0.03 | <0.001 | <0.001 |
| Emotional difficulties 3yrs | 30,385 | 0.02 | 0.00, 0.03 | 0.005 | 0.006 |
| Emotional difficulties 5 yrs | 21,556 | 0.03 | 0.01, 0.04 | <0.001 | <0.001 |
| Behavioral difficulties 1.5yrs | 39,347 | 0.01 | 0.00, 0.02 | 0.004 | 0.005 |
| Behavioral difficulties 3yrs | 30,391 | 0.03 | 0.02, 0.04 | <0.001 | <0.001 |
| Behavioral difficulties 5 yrs | 21,556 | 0.05 | 0.03, 0.06 | <0.001 | <0.001 |
| Depressive symptoms 8yrs | 22,698 | 0.06 | 0.04, 0.07 | <0.001 | <0.001 |
| Anxiety symptoms 8yrs | 22,737 | 0.03 | 0.02, 0.04 | <0.001 | <0.001 |
| Inattention 8yrs | 22,722 | 0.04 | 0.02, 0.05 | <0.001 | <0.001 |
| Oppositional defiant disorder symptoms 8yrs | 22,714 | 0.05 | 0.03, 0.06 | <0.001 | <0.001 |
| Hyperactivity 8yrs | 22,718 | 0.03 | 0.02, 0.05 | <0.001 | <0.001 |
| Conduct disorder symptoms 8yrs | 22,744 | 0.04 | 0.03, 0.06 | <0.001 | <0.001 |
| ^1^CI = Confidence Interval | | | | | |
| ^2^False discovery rate correction for multiple testing using Benjamini-Hochberg false discovery rate procedure | | | | | |

Table S4: Neuroticism PGS. Linear association between standardized PGS score and standardized score of emotional and behavioral difficulties.

| Characteristic | *n* | $\beta$ | 95% CI^1^ | p-value | p-FDR^2^ |
| --- | --- | --- | --- | --- | --- |
| Emotional difficulties 1.5yrs | 36,984 | 0.04 | 0.03, 0.05 | <0.001 | <0.001 |
| Emotional difficulties 3yrs | 30,385 | 0.04 | 0.03, 0.05 | <0.001 | <0.001 |
| Emotional difficulties 5 yrs | 21,556 | 0.04 | 0.03, 0.06 | <0.001 | <0.001 |
| Behavioral difficulties 1.5yrs | 39,347 | 0.03 | 0.02, 0.04 | <0.001 | <0.001 |
| Behavioral difficulties 3yrs | 30,391 | 0.03 | 0.02, 0.04 | <0.001 | <0.001 |
| Behavioral difficulties 5 yrs | 21,556 | 0.04 | 0.03, 0.05 | <0.001 | <0.001 |
| Depressive symptoms 8yrs | 22,698 | 0.07 | 0.05, 0.08 | <0.001 | <0.001 |
| Anxiety symptoms 8yrs | 22,737 | 0.05 | 0.04, 0.07 | <0.001 | <0.001 |
| Inattention 8yrs | 22,722 | 0.03 | 0.02, 0.04 | <0.001 | <0.001 |
| Oppositional defiant disorder symptoms 8yrs | 22,714 | 0.04 | 0.03, 0.06 | <0.001 | <0.001 |
| Hyperactivity 8yrs | 22,718 | 0.02 | 0.01, 0.04 | <0.001 | <0.001 |
| Conduct disorder symptoms 8yrs | 22,744 | 0.02 | 0.00, 0.03 | 0.007 | 0.009 |
| ^1^CI = Confidence Interval | | | | | |
| ^2^False discovery rate correction for multiple testing using Benjamini-Hochberg false discovery rate procedure | | | | | |

### Table S5: Anxiety disorder PGS. Linear association between standardized PGS score and standardized score of emotional and behavioral difficulties.

| Characteristic | *n* | $\beta$ | 95% CI^1^ | p-value | p-FDR^2^ |
| --- | --- | --- | --- | --- | --- |
| Emotional difficulties 1.5yrs | 36,984 | 0.01 | 0.00, 0.02 | 0.026 | 0.033 |
| Emotional difficulties 3yrs | 30,385 | 0.02 | 0.01, 0.03 | 0.003 | 0.004 |
| Emotional difficulties 5 yrs | 21,556 | 0.03 | 0.02, 0.04 | <0.001 | <0.001 |
| Behavioral difficulties 1.5yrs | 39,347 | 0.01 | 0.00, 0.02 | 0.058 | 0.070 |
| Behavioral difficulties 3yrs | 30,391 | 0.02 | 0.01, 0.03 | 0.004 | 0.005 |
| Behavioral difficulties 5 yrs | 21,556 | 0.03 | 0.01, 0.04 | <0.001 | <0.001 |
| Depressive symptoms 8yrs | 22,698 | 0.04 | 0.03, 0.05 | <0.001 | <0.001 |
| Anxiety symptoms 8yrs | 22,737 | 0.03 | 0.01, 0.04 | <0.001 | <0.001 |
| Inattention 8yrs | 22,722 | 0.02 | 0.01, 0.04 | <0.001 | <0.001 |
| Oppositional defiant disorder symptoms 8yrs | 22,714 | 0.03 | 0.01, 0.04 | <0.001 | <0.001 |
| Hyperactivity 8yrs | 22,718 | 0.02 | 0.01, 0.04 | <0.001 | <0.001 |
| Conduct disorder symptoms 8yrs | 22,744 | 0.02 | 0.01, 0.03 | 0.003 | 0.004 |
| ^1^CI = Confidence Interval | | | | | |
| ^2^False discovery rate correction for multiple testing using Benjamini-Hochberg false discovery rate procedure | | | | | |

Table S6: Bipolar disorder PGS. Linear association between standardized PGS score and standardized score of emotional and behavioral difficulties.

| Characteristic | *n* | $\beta$ | 95% CI^1^ | p-value | p-FDR^2^ |
| --- | --- | --- | --- | --- | --- |
| Emotional difficulties 1.5yrs | 36,984 | -0.01 | -0.02, 0.00 | 0.2 | 0.3 |
| Emotional difficulties 3yrs | 30,385 | 0.00 | -0.02, 0.01 | 0.4 | 0.4 |
| Emotional difficulties 5 yrs | 21,556 | -0.01 | -0.02, 0.00 | 0.2 | 0.2 |
| Behavioral difficulties 1.5yrs | 39,347 | -0.01 | -0.02, 0.00 | 0.10 | 0.12 |
| Behavioral difficulties 3yrs | 30,391 | -0.01 | -0.02, 0.01 | 0.3 | 0.3 |
| Behavioral difficulties 5 yrs | 21,556 | 0.00 | -0.01, 0.02 | 0.6 | 0.6 |
| Depressive symptoms 8yrs | 22,698 | 0.02 | 0.01, 0.03 | 0.002 | 0.003 |
| Anxiety symptoms 8yrs | 22,737 | 0.00 | -0.01, 0.02 | 0.7 | 0.7 |
| Inattention 8yrs | 22,722 | 0.00 | -0.01, 0.02 | 0.5 | 0.5 |
| Oppositional defiant disorder symptoms 8yrs | 22,714 | 0.03 | 0.02, 0.04 | <0.001 | <0.001 |
| Hyperactivity 8yrs | 22,718 | 0.02 | 0.01, 0.03 | 0.002 | 0.004 |
| Conduct disorder symptoms 8yrs | 22,744 | 0.03 | 0.02, 0.04 | <0.001 | <0.001 |
| ^1^CI = Confidence Interval | | | | | |
| ^2^False discovery rate correction for multiple testing using Benjamini-Hochberg false discovery rate procedure | | | | | |

### Table S7 Model fit for basic linear latent growth models across early childhood

| **Measurement** | **CFI** | **TIL** | **RMSEA (95% CI)** |
| --- | --- | --- | --- |
| Emotional difficulties | 0.993 | 0.939 | 0.032 (0.025, 0.039) |
| Behavioral difficulties | 0.992 | 0.927 | 0.044 (0.037, 0.051) |

**Note:** CFI= Comparative fit index; TLI=Tucker Lewis Index and RMSEA=Root Mean Square Error of Approximation.

### Table S8 Evaluating best fitting latent growth model for emotional and behavioral difficulties for PGS of depression

| Childhood difficulty | Model | Df | AIC | Chisq.diff | P(>Chisq) |
| --- | --- | --- | --- | --- | --- |
| **Emotional difficulties** |  |  |  |  |  |
|  | Age specific PGS effects | 1 | 290246.4 |  |  |
|  | PGS effect on growth factors | 2 | 290244.7 | 0.258 | 0.611 |
|  | PGS effect on growth factors | 2 | 290244.7 |  |  |
|  | **PGS effect on intercept only** | **3** | **290243.4** | **0.650** | **0.420** |
|  | PGS effect on growth factors | 2 | 290244.7 |  |  |
|  | PGS effect on slope only | 3 | 290257.4 | 14.667 | 0.000 |
|  | PGS effect on intercept only | 3 | 290243.4 |  |  |
|  | No PGS effect | 4 | 290265.8 | 24.433 | 0.000 |
| **Behavioral difficulties** |  |  |  |  |  |
|  | Age specific PGS effects | 1 | 399123.6 |  |  |
|  | **PGS effect on growth factors** | **2** | **399122.6** | **1.008** | **0.315** |
|  | PGS effect on growth factors | 2 | 399122.6 |  |  |
|  | PGS effect on intercept only | 3 | 399143.0 | 22.380 | 0.000 |
|  | PGS effect on growth factors | 2 | 399122.6 |  |  |
|  | PGS effect on slope only | 3 | 399131.9 | 11.280 | 0.001 |
|  | PGS effect on growth factors | 2 | 399122.6 |  |  |
|  | No PGS effect | 4 | 399184.4 | 65.842 | 0.000 |

**Note:** Df= Degrees of Freedom, AIC= First-order Alkaline Information Criteria, Chisq.diff= Difference in Chi Square Value. P(>Chisq) = P-value for Chi Square Test. Finally selected model is highlighted in bold.

### Table S9 Evaluating best fitting latent growth model for emotional and behavioral difficulties for PGS of anxiety

| Childhood difficulty | Model | Df | AIC | Chisq.diff | P(>Chisq) |
| --- | --- | --- | --- | --- | --- |
| **Emotional difficulties** |  |  |  |  |  |
|  | Age specific PGS effects | 1 | 290253.3 |  |  |
|  | **PGS effect on growth factors** | **2** | **290251.3** | **0.019** | **0.889** |
|  | PGS effect on growth factors | 2 | 290251.3 |  |  |
|  | PGS effect on intercept only | 3 | 290253.4 | 4.085 | 0.043 |
|  | PGS effect on growth factors | 2 | 290251.3 |  |  |
|  | PGS effect on slope only | 3 | 290254.2 | 4.948 | 0.026 |
|  | PGS effect on growth factors | 2 | 290251.3 |  |  |
|  | No PGS effect | 4 | 290265.8 | 18.502 | 0.000 |
| **Behavioral difficulties** |  |  |  |  |  |
|  | Age specific PGS effects | 1 | 399172.7 |  |  |
|  | **PGS effect on growth factors** | **2** | **399170.9** | **0.157** | **0.692** |
|  | PGS effect on growth factors | 2 | 399170.9 |  |  |
|  | PGS effect on intercept only | 3 | 399173.2 | 4.351 | 0.037 |
|  | PGS effect on growth factors | 2 | 399170.9 |  |  |
|  | PGS effect on slope only | 3 | 399173.4 | 4.472 | 0.034 |
|  | PGS effect on growth factors | 2 | 399170.9 |  |  |
|  | No PGS effect | 4 | 399184.4 | 17.543 | 0.000 |

**Note:** Df= Degrees of Freedom, AIC= First-order Alkaline Information Criteria, Chisq.diff= Difference in Chi Square Value. P(>Chisq) = P-value for Chi Square Test. Finally selected model is highlighted in bold.

### Table S10 Evaluating best fitting latent growth model for emotional and behavioral difficulties and PGS of neuroticism

| Childhood difficulty | Model | Df | AIC | Chisq.diff | P(>Chisq) |
| --- | --- | --- | --- | --- | --- |
| **Emotional difficulties** |  |  |  |  |  |
|  | Age specific PGS effects | 1 | 290182.7 |  |  |
|  | PGS effect on growth factors | 2 | 290181.0 | 0.364 | 0.546 |
|  | PGS effect on growth factors | 2 | 290181.0 |  |  |
|  | **PGS effect on intercept only** | **3** | **290180.9** | **1.921** | **0.166** |
|  | PGS effect on growth factors | 2 | 290181.0 |  |  |
|  | PGS effect on slope only | 3 | 290232.3 | 53.270 | 0.000 |
|  | PGS effect on intercept only | 3 | 290180.9 |  |  |
|  | No PGS effect | 4 | 290265.8 | 86.841 | 0.000 |
| **Behavioral difficulties** |  |  |  |  |  |
|  | Age specific PGS effects | 1 | 399119.8 |  |  |
|  | PGS effect on growth factors | 2 | 399117.8 | 0.042 | 0.837 |
|  | PGS effect on growth factors | 2 | 399117.8 |  |  |
|  | **PGS effect on intercept only** | **3** | **399117.9** | **2.109** | **0.146** |
|  | PGS effect on growth factors | 2 | 399117.8 |  |  |
|  | PGS effect on slope only | 3 | 399157.6 | 41.791 | 0.000 |
|  | PGS effect on intercept only | 3 | 399117.9 |  |  |
|  | No PGS effect | 4 | 399184.4 | 68.494 | 0.000 |

**Note:** Df= Degrees of Freedom, AIC= First-order Alkaline Information Criteria, Chisq.diff= Difference in Chi Square Value. P(>Chisq) = P-value for Chi Square Test. Finally selected model is highlighted in bold.

### Table S11 Evaluating best fitting latent growth model for emotional and behavioral difficulties for PGS of bipolar disorder

| Childhood difficulty | Model | Df | AIC | Chisq.diff | P(>Chisq) |
| --- | --- | --- | --- | --- | --- |
| **Emotional difficulties** |  |  |  |  |  |
|  | Age specific PGS effects | 1 | 290269.9 |  |  |
|  | PGS effect on growth factors | 2 | 290267.9 | 0.023 | 0.880 |
|  | PGS effect on growth factors | 2 | 290267.9 |  |  |
|  | PGS effect on intercept only | 3 | 290266.0 | 0.080 | 0.777 |
|  | PGS effect on growth factors | 2 | 290267.9 |  |  |
|  | PGS effect on slope only | 3 | 290266.9 | 1.009 | 0.315 |
|  | PGS effect on intercept only | 3 | 290266.0 |  |  |
|  | **No PGS effect** | **4** | **290265.8** | **1.790** | **0.181** |
| **Behavioral difficulties** |  |  |  |  |  |
|  | Age specific PGS effects | 1 | 399186.0 |  |  |
|  | PGS effect on growth factors | 2 | 399184.3 | 0.338 | 0.561 |
|  | PGS effect on growth factors | 2 | 399184.3 |  |  |
|  | PGS effect on intercept only | 3 | 399184.3 | 1.969 | 0.161 |
|  | PGS effect on growth factors | 2 | 399184.3 |  |  |
|  | PGS effect on slope only | 3 | 399186.2 | 3.879 | 0.049 |
|  | PGS effect on intercept only | 3 | 399184.3 |  |  |
|  | **No PGS effect** | **4** | **399184.4** | **2.149** | **0.143** |

**Note:** Df= Degrees of Freedom, AIC= First-order Alkaline Information Criteria, Chisq.diff= Difference in Chi Square Value. P(>Chisq) = P-value for Chi Square Test. Finally selected model is highlighted in bold.

### Table S12: Model fit statistics for latent profile analyses

| **Model** | **LL** | **AIC** | **AICc** | **Entropy** | **VLMR 2LL Diff** | **VLMR test *p* value** |
| --- | --- | --- | --- | --- | --- | --- |
| 1 profile | -639955.21 | 1280004.41 | 1280004.53 |  |  |  |
| 2 profiles | -632923.412 | 1265962.82 | 1265962.98 | 0.815 | 14063.59 | <0.001 |
| 3 profiles | -629359.153 | 1258856.31 | 1258856.53 | 0.786 | 7128.52 | <0.001 |
| 4 profiles | -625839.72 | 1251839.44 | 1251839.74 | 0.761 | 6283.80 | <0.001 |
| 5 profiles | -623605.72 | 1247393.44 | 1247393.82 | 0.759 | 4468.00 | 0.0024 |
| 6 profiles | -621977.53 | 1244159.05 | 1244159.54 | 0.762 | 2230.48 | 0.3189 |
| 7 profiles | -620662.17 | 1241550.35 | 1241550.94 | 0.749 | 1231.34 | 0.1393 |
| 8 profiles | -618652.72 | 1237553.43 | 1237554.15 | 0.759 | 2639.33 | 0.1864 |

**Note:** LL= Loglikelihood value for final model. AIC= First-order Akaike Information Criterion. AICc= Second-order Akaike Information Criterion. VLMR= Vuong–Lo–Mendell–Rubin likelihood ratio test.

### Table S13: Distribution of individuals in each profile with five-profile-model

| **profile** | **count** | **proportion** | **prob_Class**  **1** | **prob_Clas**  **2** | **prob_Class3** | **prob_Class4** | **prob_Class5** |
| --- | --- | --- | --- | --- | --- | --- | --- |
| 1 | 2099 | 0.04815 | 0.784 | 0.141 | 0.023 | 0.039 | 0.013 |
| 2 | 37071 | 0.85041 | 0.028 | 0.878 | 0.057 | 0.028 | 0.01 |
| 3 | 2113 | 0.04847 | 0.005 | 0.06 | 0.912 | 0.016 | 0.007 |
| 4 | 1828 | 0.04193 | 0.027 | 0.114 | 0.078 | 0.703 | 0.077 |
| 5 | 481 | 0.01103 | 0 | 0 | 0.03 | 0.004 | 0.966 |

Table S14: Relative odds of any emotional disorder given assignment to specific developmental profile.

| **Comparison** | **Odds ratio (OR)** | **95% CI** | |
| --- | --- | --- | --- |
|  |  | **Lower 2.5%** | **Upper 2.5%** |
| Profile 1 vs. Reference | 2.800 | 2.351 | 3.249 |
| Profile 3 vs. Reference | 1.534 | 1.248 | 1.820 |
| Profile 4 vs. Reference | 1.804 | 1.390 | 2.218 |
| Profile 5 vs. Reference | 2.937 | 2.128 | 3.746 |
| Profile 3 vs. Profile 1 | 0.548 | 0.425 | 0.671 |
| Profile 4 vs. Profile 1 | 0.644 | 0.470 | 0.818 |
| Profile 5 vs. Profile 1 | 1.049 | 0.727 | 1.371 |
| Profile 4 vs. Profile 3 | 1.176 | 0.841 | 1.511 |
| Profile 5 vs. Profile 3 | 1.914 | 1.313 | 2.516 |
| Profile 5 vs. Profile 4 | 1.628 | 1.020 | 2.235 |

**Note:** All profiles were compared to each other.

### Table S15: Relative odds of assignment to specific developmental profile per standard deviation increase in polygenic score for depression.

| **Comparison** | **Odds ratio (OR)** | **95% CI** | |
| --- | --- | --- | --- |
|  |  | **Lower 2.5%** | **Upper 2.5%** |
| Profile 1 vs. Reference | 1.130 | 1.063 | 1.200 |
| Profile 3 vs. Reference | 1.112 | 1.053 | 1.174 |
| Profile 4 vs. Reference | 1.153 | 1.067 | 1.246 |
| Profile 5 vs. Reference | 1.217 | 1.096 | 1.351 |
| Profile 3 vs. Profile 1 | 0.984 | 0.912 | 1.062 |
| Profile 4 vs. Profile 1 | 1.021 | 0.925 | 1.126 |
| Profile 5 vs. Profile 1 | 1.077 | 0.957 | 1.213 |
| Profile 4 vs. Profile 3 | 1.037 | 0.944 | 1.139 |
| Profile 5 vs. Profile 3 | 1.094 | 0.976 | 1.227 |
| Profile 5 vs. Profile 4 | 1.056 | 0.918 | 1.214 |

**Note**: All profiles were compared to each other.

### Table S16: Relative odds of assignment to specific developmental profile per standard deviation increase in polygenic score for anxiety.

| **Comparison** | **Odds ratio (OR)** | **95% CI** | |
| --- | --- | --- | --- |
|  |  | **Lower 2.5%** | **Upper 2.5%** |
| Profile 1 vs. Reference | 1.109 | 1.047 | 1.175 |
| Profile 3 vs. Reference | 1.064 | 1.008 | 1.122 |
| Profile 4 vs. Reference | 1.048 | 0.974 | 1.128 |
| Profile 5 vs. Reference | 1.089 | 0.985 | 1.204 |
| Profile 3 vs. Profile 1 | 0.959 | 0.891 | 1.032 |
| Profile 4 vs. Profile 1 | 0.945 | 0.861 | 1.036 |
| Profile 5 vs. Profile 1 | 0.982 | 0.876 | 1.100 |
| Profile 4 vs. Profile 3 | 0.985 | 0.901 | 1.077 |
| Profile 5 vs. Profile 3 | 1.024 | 0.916 | 1.143 |
| Profile 5 vs. Profile 4 | 1.039 | 0.911 | 1.185 |

**Note:** All profiles were compared to each other.

### Table S17: Relative odds of assignment to specific developmental profile per standard deviation increase in polygenic score for bipolar disorder.

| **Comparison** | **Odds ratio (OR)** | **95% CI** | |
| --- | --- | --- | --- |
|  |  | **Lower 2.5%** | **Upper 2.5%** |
| Profile 1 vs. Reference | 1.011 | 0.954 | 1.072 |
| Profile 3 vs. Reference | 1.039 | 0.985 | 1.096 |
| Profile 4 vs. Reference | 1.001 | 0.929 | 1.079 |
| Profile 5 vs. Reference | 1.198 | 1.082 | 1.327 |
| Profile 3 vs. Profile 1 | 1.027 | 0.954 | 1.107 |
| Profile 4 vs. Profile 1 | 0.990 | 0.901 | 1.088 |
| Profile 5 vs. Profile 1 | 1.185 | 1.056 | 1.330 |
| Profile 4 vs. Profile 3 | 0.964 | 0.880 | 1.055 |
| Profile 5 vs. Profile 3 | 1.154 | 1.031 | 1.290 |
| Profile 5 vs. Profile 4 | 1.197 | 1.047 | 1.369 |

**Note**: All profiles were compared to each other.

### Table S18: Relative odds of assignment to specific developmental profile per standard deviation increase in polygenic score for neuroticism.

| **Comparison** | **Odds ratio (OR)** | **95% CI** | |
| --- | --- | --- | --- |
|  |  | **Lower 2.5%** | **Upper 2.5%** |
| Profile 1 vs. Reference | 1.209 | 1.139 | 1.283 |
| Profile 3 vs. Reference | 1.060 | 1.004 | 1.118 |
| Profile 4 vs. Reference | 1.157 | 1.075 | 1.245 |
| Profile 5 vs. Reference | 1.056 | 0.952 | 1.170 |
| Profile 3 vs. Profile 1 | 0.877 | 0.813 | 0.945 |
| Profile 4 vs. Profile 1 | 0.957 | 0.871 | 1.051 |
| Profile 5 vs. Profile 1 | 0.873 | 0.777 | 0.982 |
| Profile 4 vs. Profile 3 | 1.091 | 0.998 | 1.193 |
| Profile 5 vs. Profile 3 | 0.996 | 0.890 | 1.115 |
| Profile 5 vs. Profile 4 | 0.913 | 0.799 | 1.043 |

**Note:** All profiles were compared to each other.
